# Unveiling Sex-based Differences in Parkinson’s Disease: A Comprehensive Meta-analysis of Transcriptomic Studies

**DOI:** 10.1101/2021.10.22.21265376

**Authors:** Adolfo López-Cerdán, Zoraida Andreu, Marta R. Hidalgo, Rubén Grillo-Risco, José Francisco Català-Senent, Irene Soler-Sáez, Almudena Neva-Alejo, Fernando Gordillo, María de la Iglesia-Vayá, Francisco García-García

## Abstract

In recent decades, increasing longevity (among other factors) has fostered a rise in Parkinson’s disease (PD) incidence. Although not exhaustively studied in this devastating disease, the impact of sex represents a critical variable in PD as epidemiological and clinical features differ between males and females. To study sex bias in PD, we conducted a systematic review, tissue-specific meta-analyses of transcriptomic data from the frontal cortex (FC), striatum tissue (ST), and substantia nigra (SN), and a global meta-analysis of all 3 brain regions. We selected 7 PD studies that included sex information, analyzing 267 samples. The tissue-specific meta-analyses linked PD to the enhanced expression of *MED31* in the female FC and the dysregulation of 237 genes in the SN. The global meta-analysis detected 15 genes with sex-differential patterns in PD, which participate in mitochondrial function, oxidative stress, neuronal degeneration, and cell death. Furthermore, functional analyses identified pathways, protein-protein interaction networks, and transcription factors that differed by sex. While male PD patients exhibited changes in oxidative stress based on metal ions, inflammation, and angiogenesis, female PD patients exhibited dysfunctions in mitochondrial and lysosomal activity, antigen processing and presentation functions, and glutamic and purine metabolism. Overall, we believe that our findings will provide new insight into the perspective of sex in PD, thereby underscoring the importance of including sex information in future PD studies.

## INTRODUCTION

Parkinson’s disease (PD) is the second most common progressive neurodegenerative disease after Alzheimer’s disease and the most rapidly-expanding related disorder in the elderly population, affecting 1% of individuals over 65 and 2% of individuals over 85 [1]. Importantly, PD also occurs in younger patients, with a prevalence of 0.004% in individuals between the ages of 40 and 50 [2]. PD affects different areas of the brain, with the frontal cortex (FC - involved in mental tasks), the striatum tissue (ST - coordinates multiple aspects of cognition including motor and action planning, decision-making, motivation, reinforcement, and reward perception), and the substantia nigra (SN - controls eye movement, motor planning, reward-seeking, learning, and addiction) the most affected. The selective degeneration of SN dopaminergic neurons that innervate the ST (nigrostriatal dopaminergic neurons - NSDA) and the formation of Lewy bodies (α-synuclein accumulation in neurons) in several different brain regions [3] prompt motor symptoms (e.g., resting tremor, muscle stiffness, bradykinesia, and postural instability) and non-motor symptoms such as cognitive and mood impairment. In advanced-stage PD, a third of patients develop dementia [4]. Oxidative stress, excitotoxicity, and neuroinflammation influence neuron death in PD [5,6], while nitric oxide and other reactive nitrogen species [7,8] promote PD progression. Mitochondrial dysfunction also plays a significant role in PD, as in other neurodegenerative diseases [9]. While the etiology of PD remains unclear, studies have provided evidence that genetic factors and environmental triggers, including genetic variation, sex, age, and exposure to environmental toxins (e.g., herbicides and pesticides), all play roles in pathology development [10].

Importantly, male and female PD patients display specific sex-based differences [11]. While male PD patients suffer from a greater risk of PD [12], female PD patients suffer from higher mortality rates and require earlier documented placements in nursing care [13]. Other sex-based differences in PD include nigrostriatal degeneration, time of symptom onset, motor and non-motor symptoms, REM sleep behavior disorder, treatment outcomes, and disease mechanisms [14–18]. Inflammation and mitochondrial function also display sex-based alterations in PD patients [19,20]. Genetic factors, sex chromosome genes, hormones, and neuroactive steroids represent the leading causes for these sex-based differences [15,18,21].

While a minority of PD cases benefit from genetic testing, routine clinical methods for PD diagnosis at early disease stages do not exist, complicating disease management. There exist therapeutic options such as l-DOPA (levodopa or l-3,4-dihydroxyphenylalanine) and deep brain stimulation, which improve symptoms and patient quality-of-life; however, these treatments do not delay neurodegeneration. Thus, identifying PD biomarkers represents an unmet clinical task that may allow for early diagnosis and the creation of improved therapies.

To evaluate sex-based differences in PD, we now present a systematic review and 4 transcriptomic meta-analyses on different brain regions followed by a functional characterization. These results demonstrate significant differences in gene expression and biological functions when comparing male and female PD patients and brain regions.

## METHODS

All bioinformatics and statistical analysis were performed using R software v.3.6.3 [22]. Additional file 2: Table S2 details R packages and versions.

### Study Search and Selection

Available datasets were collected from the Gene Expression Omnibus (GEO) [23] and ArrayExpress [24] public repositories. A systematic search of all published studies in public repositories (2002-2020) was conducted between June and September of 2020, following the preferred reporting items for systematic reviews and meta-analyses (PRISMA) guidelines [25]. Keywords employed in the search were “Parkinson”, “Parkinson’s Disease”, and “PD”. We applied the following inclusion criteria:

- Transcriptomics studies on *Homo sapiens*
- Control and PD-affected subjects included
- Sex, disease status, and brain region variables registered
- RNA extracted directly from post-mortem brain tissues (no cell lines or cultures)
- Brain tissues from either FC, ST, or SN
- Sample size > 3 for case and control groups in both sexes

Finally, normalized gene expression data of 8 array PD datasets (E-MTAB-1194, E-MEXP-1416, GSE28894, GSE8397, GSE20295, GSE20159, GSE7621, and GSE20146) were retrieved using R packages GEOquery [26] and ArrayExpress [27].

### Individual Transcriptomics Analysis

Individual transcriptomics analysis was performed on every selected study, which comprised 2 steps: preprocessing and differential expression analysis.

Data preprocessing included the standardization of the nomenclature for the clinical variables in each study, the homogenization of gene annotation, and an exploratory data analysis. The normalization methods performed by the original authors were assessed for each dataset, and data matrices were log_2_ transformed when necessary. All probe sets were annotated to HUGO gene symbols [28] using the biomaRt R package [29]. When dealing with duplicated probe-to-symbol mappings, the median of their expression values was calculated. The exploratory analysis included unsupervised clustering and PCA to detect expression patterns between samples and genes and the presence of batch effects in each study. At this point, the GSE20159 study was excluded for presenting a strong batch effect with a critical impact on differential expression analysis.

Differential expression analyses were performed using the limma R package [30] to detect sex-based differentially expressed genes. To achieve this goal, the following comparison was applied:

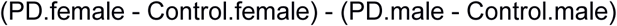

This comparison allows the detection of genes with sex-based differential behavior in the development of PD. Genes with an Log_2_ Fold Change (LFC) greater than 0 show either a higher increase or a lesser decrease in expression in females when comparing the effect of the disease between sexes. For simplicity, these genes are referred to as increased in females. On the contrary, genes with an LFC lower than 0 have a higher increase or a lesser decrease in expression in males when comparing the effect of the disease between sexes. For simplicity, these genes are referred to as increased in males.

This comparison was applied to each brain region separately and all 3 regions together. When necessary, the batch effect was included as a categorical variable on the limma linear model to reduce its impact on data. P-values were corrected using the Benjamini-Hochberg procedure [31] and considered significant below a threshold of 0.05.

### Gene Expression Meta-Analysis

Differential gene expression results were integrated into a single meta-analysis [32] for each brain region (FC, ST, and SN) and a fourth meta-analysis for all regions. Meta-analyses were implemented with the R package metafor [33], under the DerSimonian & Laird random-effects model [34], considering individual study heterogeneity. This model considers the variability of individual studies by increasing the weights of studies with less variability when computing meta-analysis results. Thus, the most robust functions between studies are highlighted.

P-values, corrected p-values, LFC, LFC standard error (SE), and LFC 95% confidence intervals (CI) were calculated for each evaluated gene. Functions and pathways with corrected p-values of < 0.05 were considered significant, and both funnel and forest plots were computed for each. These representations were evaluated to assess for possible biased results, where LFC represents the effect size of a gene, and the SE of the LFC serves as a study precision measure [35]. Sensitivity analysis (leave-one-out cross-validation) was conducted for each significant gene to verify alterations in the results due to the inclusion of any study. The Open Targets platform [36] (release 21.06) was used to explore the associations of the significant genes to PD.

### Sex-based Functional Signature in the SN

Gene meta-analysis of SN data revealed sets of differentially expressed genes between male and female PD patients. Several analyses were carried out to identify the functional implications of these differences.

Over-Representation Analysis (ORA) [37] through the R package clusterProfiler [38] was first used to determine the biological functions and pathways overrepresented in the following gene sets: i) differentially expressed “LFC > 0” genes ii) differentially expressed “LFC < 0” genes, and iii) all differentially expressed genes. P-values and corrected p-values were calculated for each GO term from the 3 GO ontologies [39] and each KEGG pathway [40]. Every function and pathway with a corrected p-value < 0.05 was labeled as over-represented in each gene set.

Protein-protein interaction (PPI) networks were then calculated using the STRING web tool for each subset of genes [41]. The total number of edges was examined, and PPI enrichment was assessed using the default parameters for each network.

Finally, a VIPER [42] analysis with human regulons obtained from the DoRothEA R package [43] was performed to estimate transcription factor activity using the consensus LFC of each gene evaluated in meta-analysis as gene expression signature. Regulons with a confidence level of A, B, C, or D were selected, excluding those with less than 25 genes (n = 217). The p-values were corrected with the Benjamini & Hochberg method. Normalized enrichment scores (NES) were calculated by VIPER as a measure of relative transcription factor activity.

### Metafun-PD Web Tool

All data and results generated in the different steps of the meta-analysis are available in the Metafun-PD web tool [44], which is freely accessible to any user and allows the confirmation of the results described in this manuscript and the exploration of other results of interest. The front-end was developed using the Angular Framework, the interactive graphics used in this web resource have been implemented with plotly [45], and the exploratory analysis cluster plot was generated with the ggplot2 R package [46].

This easy-to-use resource is divided into 7 sections: 1) summary of analysis results in each phase. Then, for each of the studies, the detailed results of the 2) exploratory analysis and 3) differential expression. 4) The gene meta-analysis results of the four different performed meta-analyses. The user can interact with the web tool through graphics and tables and search for specific information for a gene or function. Finally, sections 5-7) provide the detailed tables and figures corresponding to the results of the 3 functional profiling methods (ORA, PPI, and VIPER).

## RESULTS

To evaluate sex-based differences in PD, we performed a systematic review and 4 meta-analyses of transcriptomic studies that included information on patient sex from the GEO [23] and ArrayExpress databases [24]. The meta-analyses comprised 1 for each of the primary brain regions affected by PD pathogenesis - FC (3 studies), ST (3 studies), and SN (f studies) - and a fourth global meta-analysis for the 3 brain regions combined (7 studies). Finally, we explored the biological implications of the SN meta-analysis results by applying 3 different functional profiling methods: over-representation analysis (ORA), the generation of protein-protein interactions (PPI) networks, and the transcription factors activity analysis (Figure 1).

**Figure 1.**
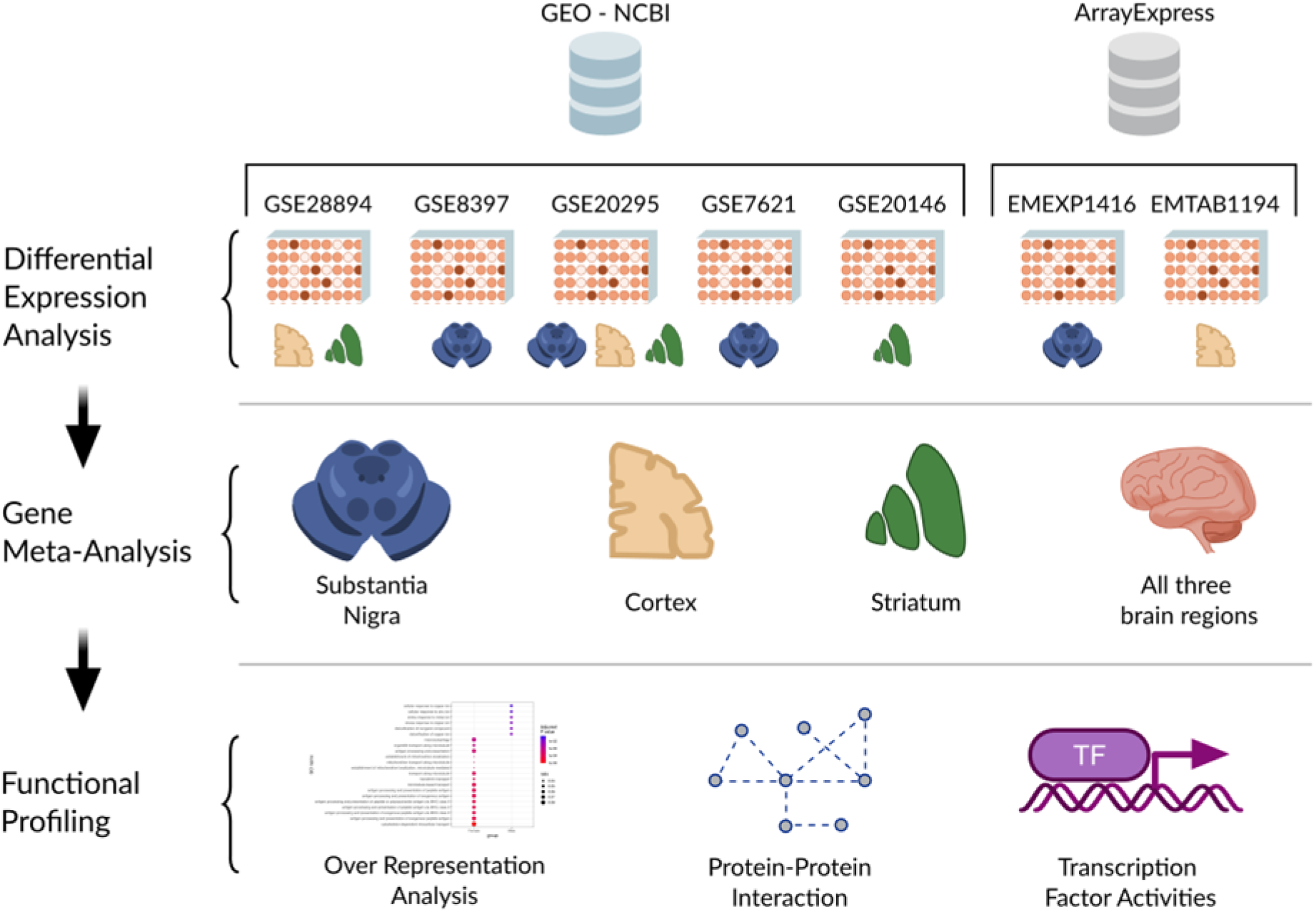
Workflow and analysis design. We retrieved relevant studies from GEO-NCBI and ArrayExpress data repositories and performed differential expression analysis on each selected study after data exploration and preprocessing. We performed a comparison for each brain region and an additional comparison for all samples and performed a gene meta-analysis for each comparison. Finally, we applied several functional profiling methodologies to characterize the results of the SN meta-analysis.

### Study Search and Selection

The systematic review identified 83 non-duplicated studies, of which 39 (47%) included both male and female PD patients. We selected 8 comparable studies after applying inclusion and exclusion criteria (Methods, Figure 2); however, we discarded 1 study after the exploratory analysis. Thus, we analyzed 7 studies that included 267 samples (132 controls and 135 PD cases) from the FC, ST, and SN brain regions (Table 1). Figure 3 describes sex distribution by study and region (overall, 59% males and 41% females); the median age was 75. Table 1 and Figure 2 contain further information regarding the selected studies and their sample clinicopathological characteristics.

**Table 1.** Studies selected after the systematic review.

| Study | Platform | Brain area | Publication |
| --- | --- | --- | --- |
| E-MTAB-1194 | Affymetrix GeneChip Human Gene 1.1 ST Array | FC | [47] |
| E-MEXP-1416 | Affymetrix GeneChip Human X3P Array | SN | [48] |
| GSE8397 | Affymetrix Human Genome U133A Array | SN | [49,50] |
| GSE20295 | Affymetrix Human Genome U133A Array | FC, SN, ST | [51,52] |
| GSE7621 | Affymetrix Human Genome U133 Plus 2.0 Array | SN | [53] |
| GSE20146 | Affymetrix Human Genome U133 Plus 2.0 Array | ST | [52] |
| GSE28894 | Illumina HumanRef-8 v2.0 Expression Beadchip | FC, ST | - |

Brain areas: cortex frontal (FC), striatum tissue (ST), and substantia nigra (SN)

**Figure 2.**
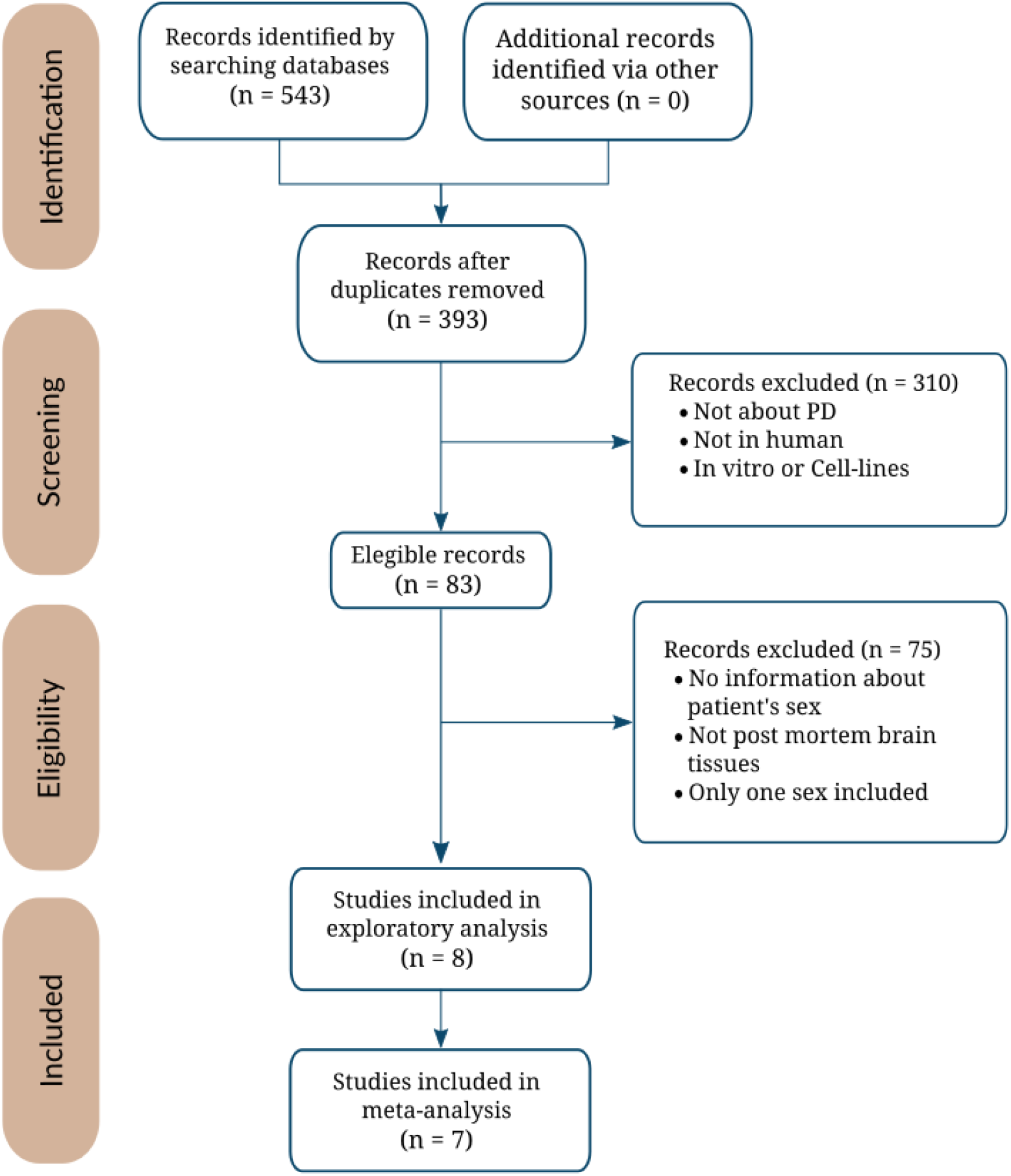
The flow of information through the distinct phases of the systematic review following PRISMA statement guidelines [25].

**Figure 3.**
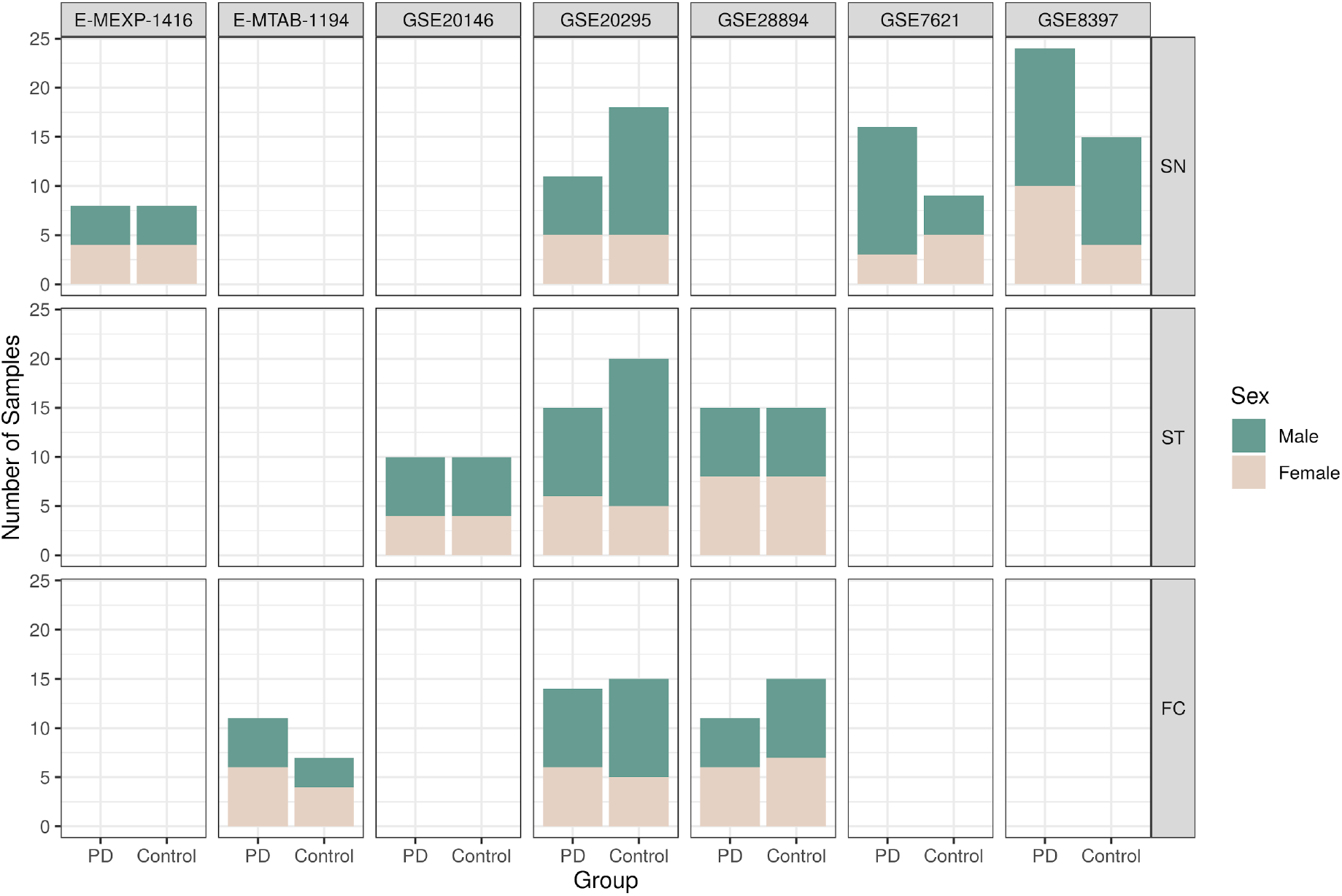
The number of samples per study, divided by sex, study, and experimental group (PD – Parkinson’s disease).

### Individual Analysis

We carried out exploratory and processing steps on the datasets to ensure their comparability in subsequent analyses. We applied log_2_ transformation to studies GSE28894, GSE20295, GSE7621, and GSE20146 to homogenize magnitude order and filtered out samples from regions different to FC, SN, and ST from all studies. Exploratory analysis revealed an anomalous behavior (outlier) by the A04022 sample from the E-MTAB-1194 study, which we excluded from further analysis.

The differential expression results for each study provided only a small number of significantly affected genes in the SN tissues from the GSE8397 study (Table 2).

**Table 2.** Summary of differential gene expression analysis by brain region using a sex-based comparison.

|  | Significant Genes |  |  | Significant Genes |  |  | Significant Genes |  |  | Significant Genes All Tissues |  |  |
| --- | --- | --- | --- | --- | --- | --- | --- | --- | --- | --- | --- | --- |
|  | FC |  |  | ST |  |  | SN |  |  |  |  |  |
|  | LFC > 0 | LFC < 0 | Total | LFC > 0 | LFC < 0 | Total | LFC > 0 | LFC < 0 | Total | LFC > 0 | LFC < 0 | Total |
| E-MTAB-1194 | 0 | 0 | 0 | - | - | - | - | - | - | 0 | 0 | 0 |
| E-MEXP-1416 | - | - | - | - | - | - | 0 | 0 | 0 | 0 | 0 | 0 |
| GSE8397 | - | - | - | - | - | - | 30 | 65 | 95 | 30 | 65 | 95 |
| GSE20295 | 0 | 0 | 0 | 0 | 0 | 0 | 0 | 0 | 0 | 0 | 0 | 0 |
| GSE7621 | - | - | - | - | - | - | 0 | 0 | 0 | 0 | 0 | 0 |
| GSE20146 | - | - | - | 0 | 0 | 0 | - | - | - | 0 | 0 | 0 |
| GSE28894 | 0 | 0 | 0 | 0 | 0 | 0 | - | - | - | 2 | 0 | 2 |

"LFC > 0" columns = Differential expressed genes increased in females; "LFC < 0" columns = Differential
expressed genes increased in males. Dash "-" indicates the absence of results due to the lack of associated tissue in the study in question. FC = frontal cortex, ST = striatum tissue, and SN = substantia nigra.

### Gene Expression Meta-Analysis

We performed a gene expression meta-analysis in the FC, ST, and SN analyzing 3 studies for FC, 3 studies for ST, and 4 for SN. Finally, we performed a global meta-analysis that integrated all 7 studies from these brain regions. In the meta-analysis by regions, we found no significant genes in the FC, 1 significant gene (MED31 - mediator of RNA polymerase II transcription subunit 31) in the ST, and 237 significant genes in the SN, reflecting the well-known impact of PD in this region. Of those, 75 genes exhibited increased expression in males (LFC < 0, see Methods) and 162 in females (LFC > 0) (Table 3). 16 and 44 of these genes in male and female PD patients, respectively, had a documented association with PD in the OpenTargets [36] database.

**Table 3.**
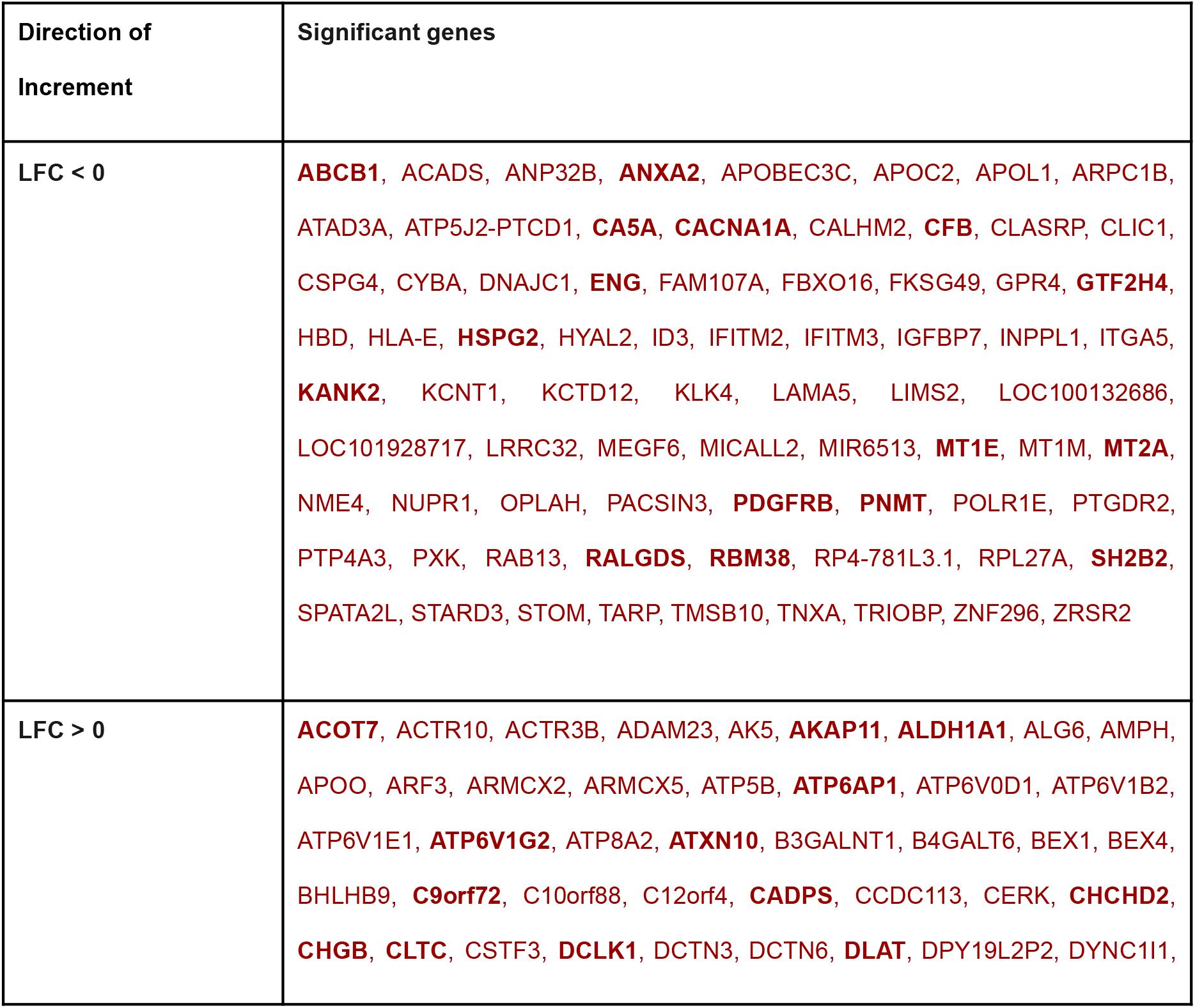
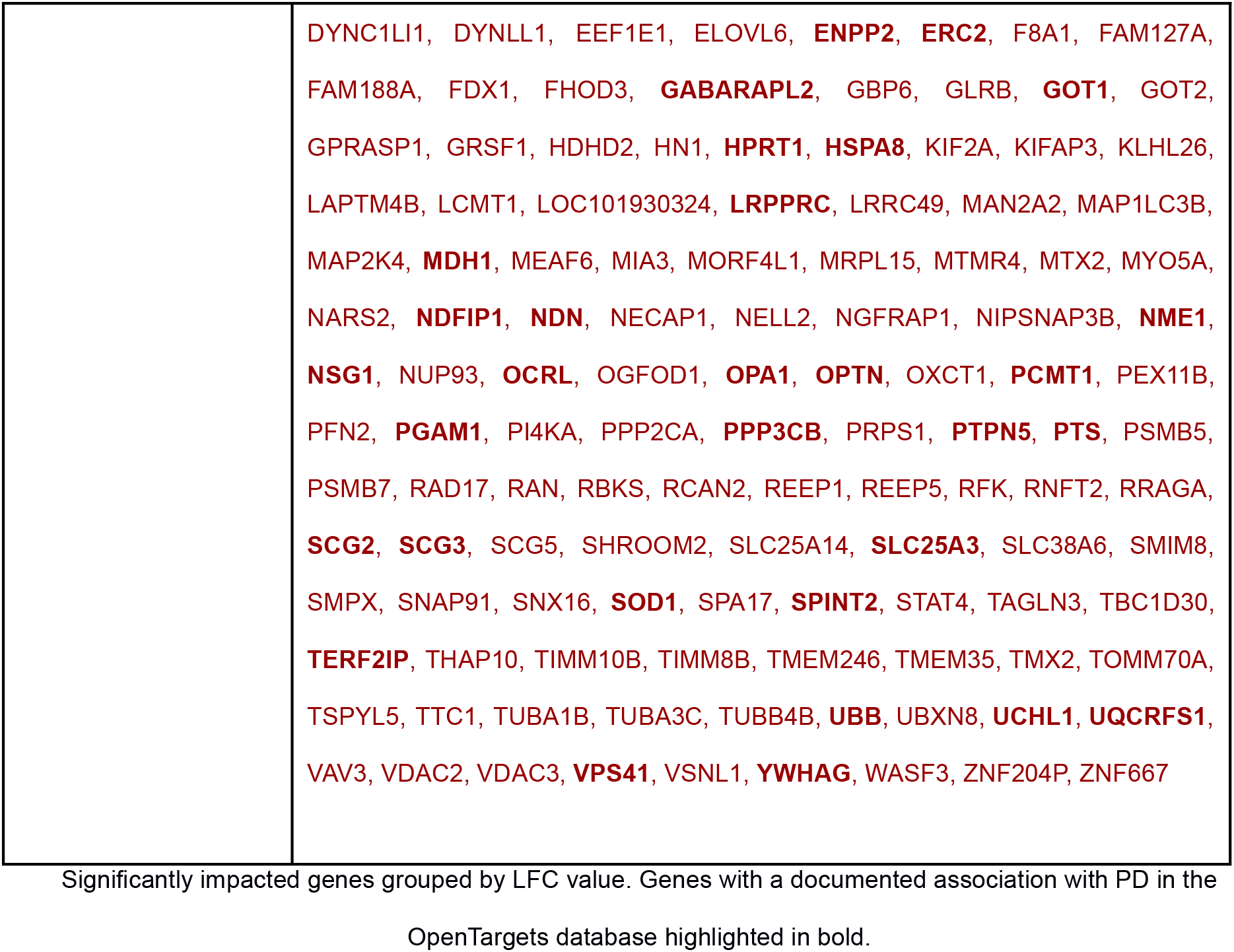
Significantly impacted genes detected in the meta-analysis of expression data from SN studies.

The global meta-analysis integrating the 7 studies and the 3 primary brain regions affected by PD revealed 15 differentially expressed genes by sex, 4 significant genes increased in females and 11 in males (Table 4). 1 gene of each group had already known associations with PD in the OpenTargets database.

**Table 4.**
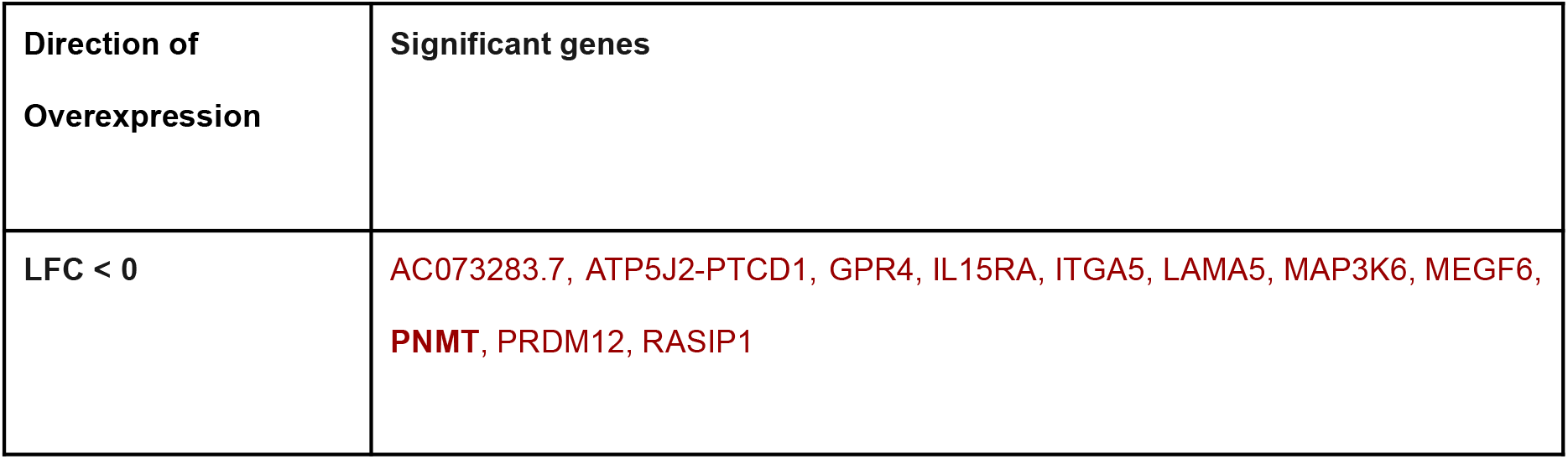
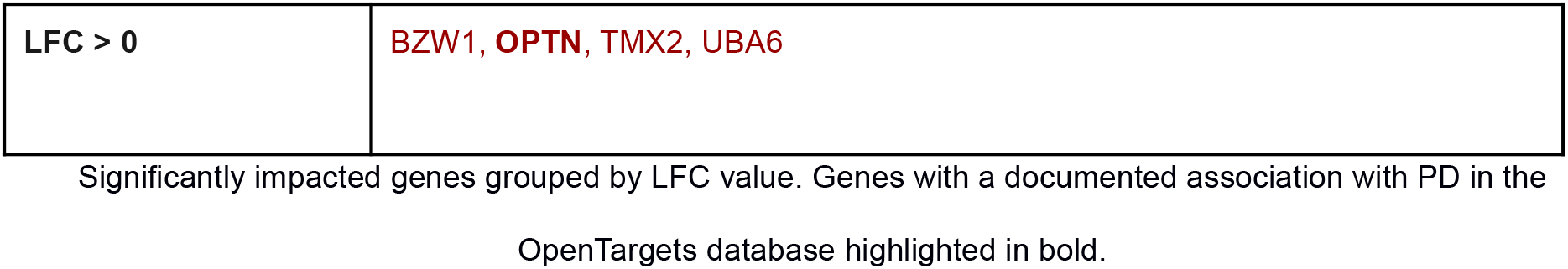
Significantly impacted genes detected in the meta-analysis of expression data for all 7 studies integrated from the 3 major brain regions.

| Direction of Overexpression | Significant genes |
| --- | --- |
| LFC < 0 | AC073283.7, ATP5J2-PTCD1, GPR4, IL15RA, ITGA5, LAMA5, MAP3K6, MEGF6, **PNMT**, PRDM12, RASIP1 |
| LFC > 0 | BZW1, **OPTN**, TMX2, UBA6 |

Significantly impacted genes grouped by LFC value. Genes with a documented association with PD in the OpenTargets database highlighted in bold.

Regarding those genes increased in males, we highlight AC073283.7 (a long non coding RNA), ATP5J2-PTCD1 (a locus that represents naturally occurring read-through transcription between the genes ATP5J2 [ATP synthase, H+ transporting, mitochondrial Fo complex, subunit F2] and PTCD1 [pentatricopeptide repeat domain 1]), GPR4 (G Protein-Coupled Receptor 4; may mediate central respiratory sensitivity to CO2 in the brain), PNMT (pentalenolactone synthase; cerebral disease and intellectual disability, hypotonia and mitochondrial disease, already linked to PD), PRDM12 (PR domain zinc finger protein 12; inflammation or degeneration of the sensory nerves), ITGA5 (Integrin Subunit Alpha 5; paraneoplastic neurologic syndrome), MEGF6 (Multiple EGF Like Domains 6 intracranial hemorrhage), LAMA5 (Laminin Subunit Alpha 5; cerebral diseases of vascular origin with epilepsy), MAP3K6 (Mitogen-Activated Protein Kinase 6, musculoskeletal or connective tissue disease genetic), IL15RA (Interleukin 15 Receptor Subunit Alpha; inflammatory brain disease), and RASIP1 (Ras Interacting Protein 1; juvenile primary lateral sclerosis [JPLS], a very rare motor neuron disease characterized by progressive upper motor neuron dysfunction leading to loss of the ability to walk; mitochondrial oxidative phosphorylation disorder with no known mechanism).

The significant genes with increased expression in females included OPTN (Optineurin; amyotrophic lateral sclerosis [ALS], a degenerative disorder affecting upper motor neurons in the brain, already linked to PD), UBA6 (Ubiquitin Like Modifier Activating Enzyme 6; developmental and epileptic encephalopathy), BZW1 (Basic Leucine Zipper And W2 Domains 1; rolandic epilepsy - speech dyspraxia; intellectual disability), and TMX2 (Thioredoxin Related Transmembrane Protein 2; regulates mitochondrial activity; abnormality in the process of thought including the ability to process information; cognitive abilities or memory anomaly, intellectual disability in general).

Additional file 1: Table S1 details the results for all significant genes from all meta-analyses, including the adjusted p-value, the LFC and its 95% confidence interval (CI), and the standard error (SE) of the LFC.

### Over-Represented Functions in the SN

ORA by R package clusterProfiler [38] on the SN significant genes revealed that enriched biological functions in the genes increased in males related to oxidative stress, including detoxification of inorganic compounds and stress response to metal ions (Figure 4). In the genes increased in females, ORA overrepresented functions relate to antigen processing and presentation, pH regulation, proton transmembrane transport, mitochondrial functions, autophagy, cytoskeleton, and microtubule transport. These functions may create a neurodegenerative microenvironment that results in neuronal death. In addition, purine metabolism and ribose metabolism also appeared as enriched in this gene set.

**Figure 4.**
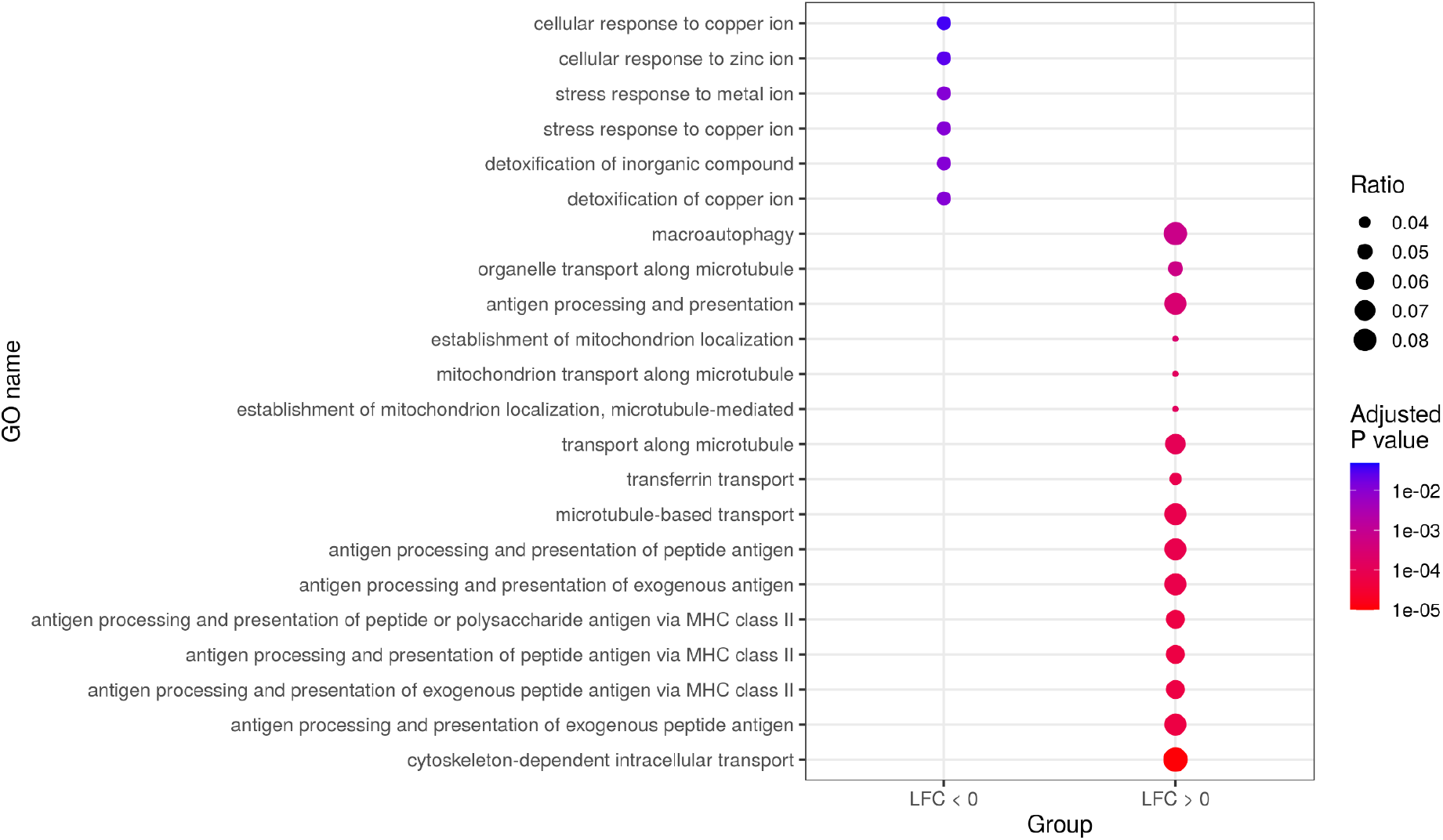
Summary dot plot of GO biological process meta-analysis results, showing only those significant functions with the highest differential effect (top 16 functions in the “LFC > 0” gene set by adjusted p-value and the 6 significant functions in the “LFC < 0” gene set). Gene ratios calculated from the subset of genes analyzed in the ORA by dividing the number of genes involved in each function by the total of genes analyzed.

### Protein-Protein Interaction Networks for SN genes

Using the STRING web tool, we created PPI networks for significant SN genes with positive and negative LFC [41]. We found high connectivity in the PPI network generated for the 75 genes with negative LFC (p-value 3.87e-05) (Figure 5a). Within the PPI analysis, we encountered the following protein clusters:

**Figure 5.**
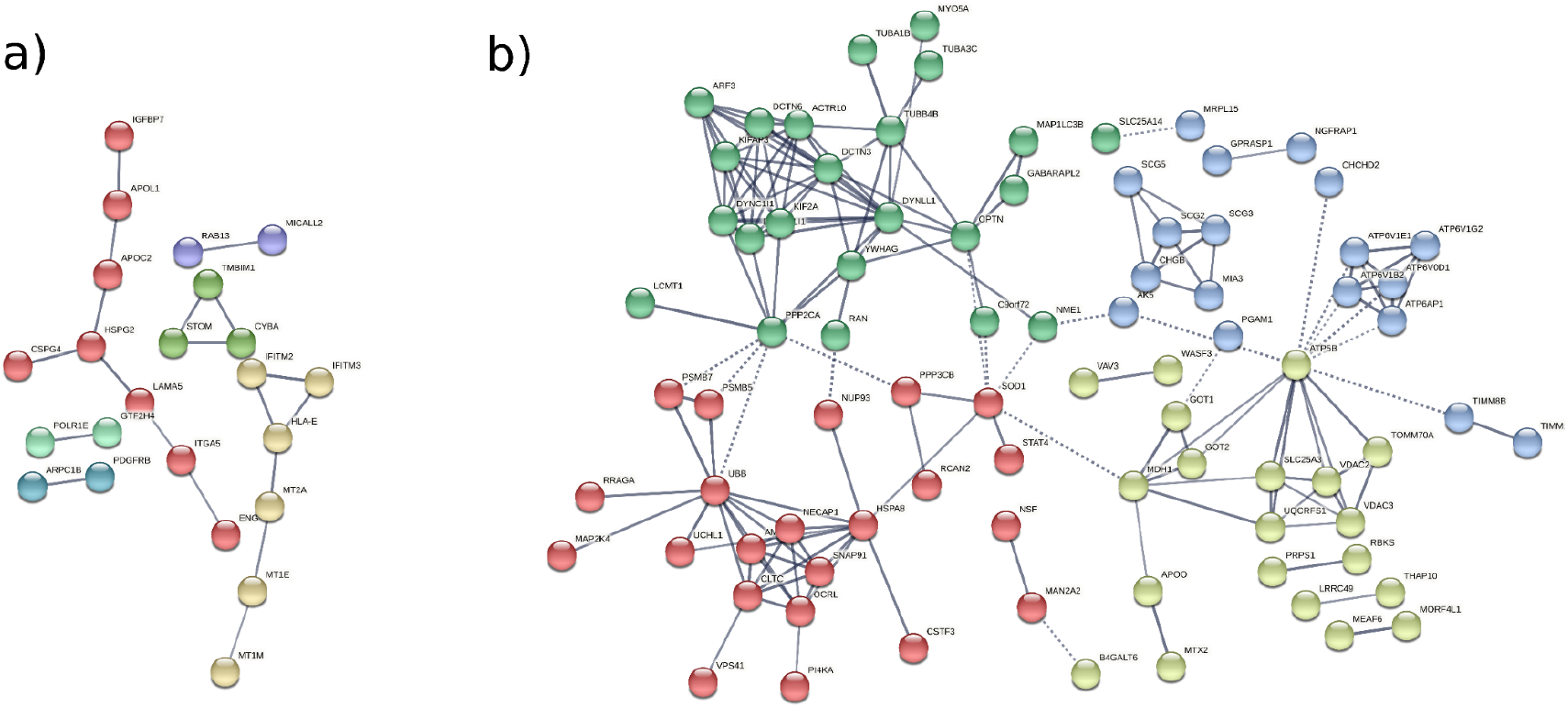
PPI networks calculated from significant genes in the SN meta-analysis, showing only network edges with an interaction score greater than 0.7. **A**) PPI network generated from the “LFC < 0” gene set. **B**) PPI network generated from the “LFC > 0” gene set.

- Angiogenesis cluster (in red): HSPG2, CSPG4, ITGA5, LAMA5, ENG, PDGFRG, APOC2, APOL1, and IGFBP7. A central cluster of proteins participates in angiogenic processes.
- Metallothioneins cluster (in pearl): MT2A, MT1E, and MT1M are linked to the interferon-induced transmembrane proteins IFITM2 and IFITM3 through HLA-E.
- STOM, TMBIM1, CYBA cluster (in green). These proteins have roles in redox homeostasis, response to oxidative stress, inflammation, and blood vessel remodeling.

We also encountered 3 independent pairs of interacting proteins: POLR1E-GTF2H4, MICALL2-RAB13, and ARPC1B-PDGFRB.

We also encountered a highly connected PPI network generated from the 162 genes with positive LFC (p-value < 1.0e-16) (Figure 5b). Within this PPI network, we encountered the following protein clusters:

- Microtubule and transport cluster (in green). This cluster includes proteins related to microtubule reorganization and transport (tubulins alpha: TUBA1B, TUBA3C, and beta TUBB4B, Myosin, and Actin).
- V-ATPase and Secretogranin clusters (in blue). Formed by V-ATPases subunits involved in neuronal diseases and members of the chromogranin/secretogranin family of neuroendocrine secretory proteins.
- Protein folding, recycling, and degradation (in red). Composed by proteasome subunits PSMB5, PSMB7; Ubiquitin UBB, Ubiquitin-protein hydrolase protein UCHL1, Chaperone HSPA8, and clathrin and adaptins (E-L system).
- Mitochondria function and metabolism cluster (in yellow). Proteins such as GOT1 and GOT2 (glutamic metabolism) or VDAC2 and VDAC3, which are voltage-dependent anion-selective channel proteins involved in mitochondrial outer membrane permeabilization, negative regulation of intrinsic apoptotic signaling pathway, and negative regulation of protein polymerization, among other functions.

### Transcription Factor Activity in the SN

The TF activity analysis of the significant SN genes performed with Dorothea [43] reported 40 differentially activated TFs (adjusted p-value < 0.05), with 29 more activated in male PD patients (NES < 0) and eleven more activated in female PD patients (NES > 0) (Figure 6 and Additional file 3: Table S3). Among the 29 TFs activated in male PD patients, ADNP, JUN, MBD3, PRDM14, and ESR1 have known associations with neurodegenerative disorders such as Alzheimer’s disease, mental retardation, schizophrenia, and autism. Additionally, a subset of specific TFs comprising BATF, CEBPA, ETS1, KLF6, LYL1, NFKB1, RELA, SOX13, SP3, SPI1, STAT1, STAT2, and STAT3 has documented involvement in immune and neuroinflammatory functions (Additional file 3: Table S3).

**Figure 6.**
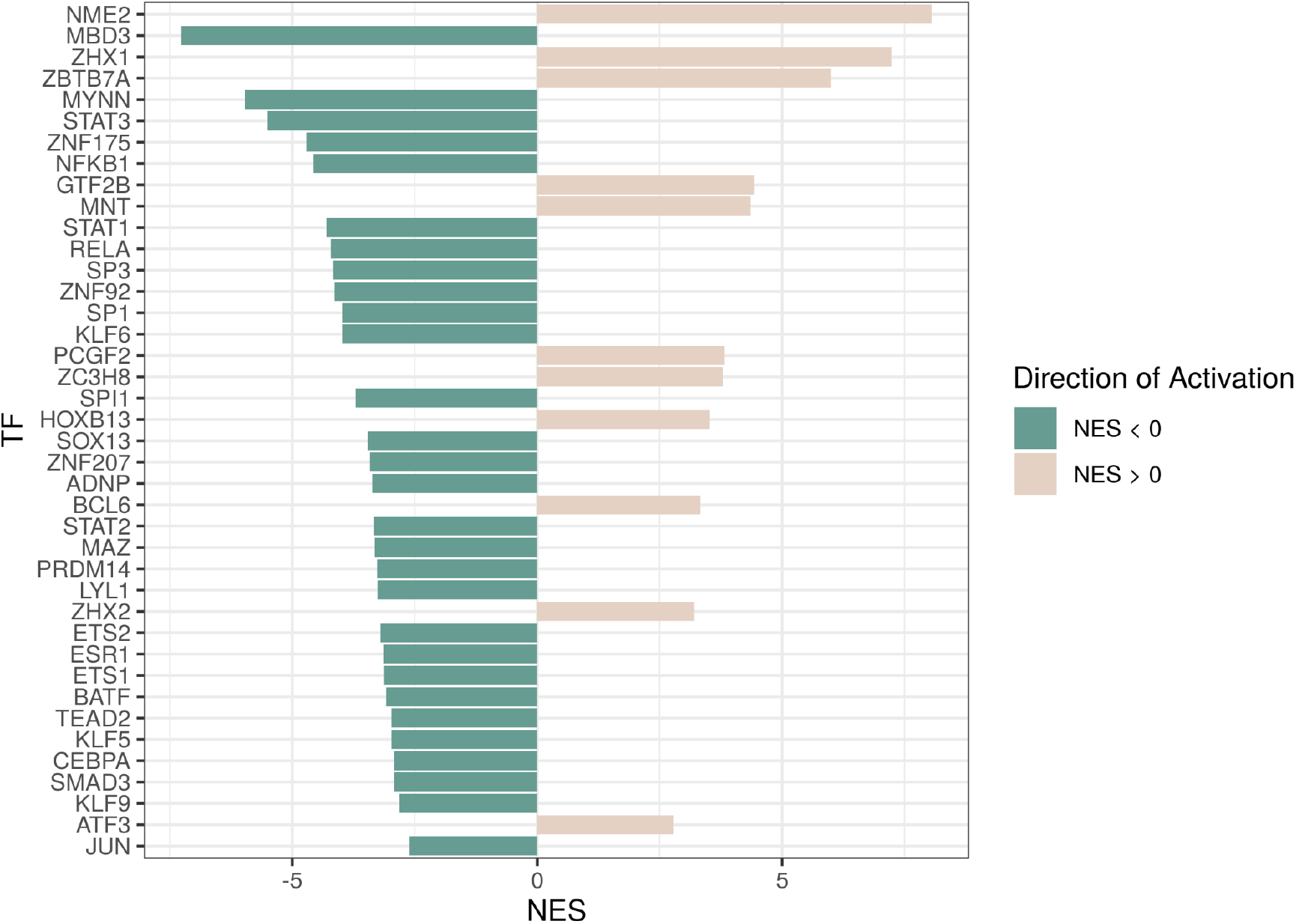
TFs with significantly altered activity (adjusted p-value < 0.05). Green indicates higher TF activity in male compared to female PD patients, while brown indicates higher TF activity in females compared to male PD patients. Activation values are measured as normalized enrichment scores (NES).

Specific TFs activated with an NES > 0 (female PD patients), such as ATF3, BCL6, or PCGF2, have been associated with neurodegenerative diseases or cognitive disabilities. TFs such as ZBTB7A, ZC3H8, or NME2 may be involved in processes related to neuroinflammation. These TFs also play additional roles in other processes, including immune response, hemopoiesis, and regulating the differentiation and activation of T and B lymphocytes (Additional file 3: Table S3). To the best of our knowledge, the remaining significant TFs remain poorly characterized or have not been studied in the context of PD.

### Metafun-PD Web Tool

The Metafun-PD web tool [44] contains information about the 7 studies and 267 samples involved in this study. This resource includes statistical indicators of each performed analysis for each study, which users can explore to identify profiles of interest.

We carried out a total of 4 meta-analyses. For each of the significantly altered genes, Metafun-PD depicts the global activation level for all studies and each study’s specific LFC, confidence interval of LFC, and p-value, and shows graphical representations by gene as forest and funnel plots. This open resource hopes to contribute to data sharing between researchers, the elaboration of innovative studies, and the discovery of new findings.

## DISCUSSION

### Meta-analysis of Transcriptomic Profiles

We carried out 3 gene expression meta-analyses in the 3 brain regions most affected by PD (FC, ST, and SN) and a fourth that integrated studies from all regions to better understand sex-based differences in PD. We identified the MED31 gene as significantly increased in females in the ST, 237 differentially expressed genes in the SN (75 and 162 significantly increased in males and females, respectively), a number that probably reflects the known impact of PD on the SN [54], but failed to find differences in the FC.

Functions associated with MED31 (mediator of RNA polymerase II transcription subunit 31) include the regulation of lipid metabolism by peroxisome proliferator-activated receptor alpha (PPARα) and gene expression [55,56]. Dysregulated lipid metabolism supports the accumulation of α-synuclein and the formation of Lewy bodies in the brainstem, limbic system, and cortical areas [57]. Long-chain fatty acids, ceramides, and lipopolysaccharides can also induce cellular stress and inflammatory responses in the brain [58]. MED31 has been associated with neural diseases such as ALS types 3 and 5 [59], which affect motor neurons; however, this study represents the first association of MED31 with PD.

The genes increased in the SN of male PD patients can be clustered into several families:

- Metallothioneins, including genes such as MT2A, MT1E, and MT1M. Metallothioneins are small cysteine-rich proteins that play essential roles in metal homeostasis and toxicity, DNA damage, and oxidative stress. Oxidative stress and inflammation (hallmarks of neurodegenerative diseases) influence the regulation of metallothionein expression. Metallothioneins have been described as potential markers of neurologic disease processes and treatment response in lysosomal storage disorders [60]. Moreover, studies have encountered increased metallothionein expression in the SN and FC of PD patients [61].
- Apolipoproteins - APOL1 and APOC2. These proteins control the binding of lipids to form lipoproteins, whose primary function involves lipid transport. Other lipoproteins, such as ApoE, have been related to aging and neurodegenerative diseases [62,63], while lipid metabolism has strong links to neurotrophic disorders and correlates with symptoms in PD patients [64].
- Interferon-induced transmembrane proteins - IFITM2 and IFITM3. These proteins play roles in the modulation of innate immunity and have been previously related to neurodegenerative diseases. In Alzheimer’s disease, inflammatory cytokines induce IFITM3 expression in neurons and astrocytes; IFTM3 then binds to γ-secretase to upregulate activity, increasing the production of amyloid-β (the main component of the amyloid plaques found in Alzheimer’s disease patients) [65,66].

In summary, male PD patients present alterations in pathways related to oxidative stress, inflammation, and innate immune response, which represent hallmarks of neurodegeneration.

Genes with a higher expression in the SN of female PD patients can also be clustered into several families:

- Secretogranins - SCG2, SCG3, SCG5, and CHGB. Members of this family control the delivery of peptides and neurotransmitters. Alterations in the granin family have been associated with PD [67].
- V-ATPase subunits - ATP5B, ATP6AP1, ATP6V0D, ATP6V1B2, ATP6V1E1, and ATP6V1G2.
- Dysfunction of V-ATPase affects lysosomal acidification, which disrupts substrate clearance and leads to many disorders, including neurodegenerative diseases [68,69].
- TIM22 complex subunits - TIMM8B and TIMM10B form a complex involved in mitochondrial protein import. Alterations in TIM complexes such as TIM23 have been suggested as relevant mechanisms in neurodegenerative diseases [70].
- Axonal transport and cytoskeleton stability: Dyneins - DYNC1LI1, DYNC1I1, and DYNLL1, Dynactin subunits - DCTN3, DCTN6, and ACTR10; Actin-related proteins - ACTR3B and ACTR10, and Tubulin subunits - TUBA1B, TUBA3C, and TUBB4B. Alterations in these complexes disrupt axonal transport, prompt the accumulation of misfolded proteins and motor neuron diseases, and are described in disorders such as ALS and other neurodegenerative processes [71–73].
- Mitochondrial porins - VDAC2 and VDAC3 are responsible for voltage-dependent anion channels and mitochondrial dysfunction and contribute to neurodegenerative diseases [74].
- Glutamic-oxaloacetic transaminase - cytoplasmic and mitochondrial forms GOT1 and GOT2. Metabolic processes also contribute to PD progression [75] and glutamate metabolism associated with excitotoxicity and neuron death [76].
- Serine/threonine phosphatase subunits - PPP3CB and PPP2CA. PP3CB is a calcium-dependent calmodulin-stimulated protein phosphatase that plays an essential role in the transduction of intracellular Ca^2+^-mediated signals [59][60]. PP3CB is related to biological processes such as axon extension, learning, locomotion, lymphangiogenesis, memory, regulation of synaptic plasticity and synaptic vesicle endocytosis, and cytokine and T cell responses. PPP2CA is the major phosphatase for microtubule-associated proteins (MAPs), which can affect GABA receptor binding, tau protein binding, regulation of apoptotic processes, and microtubule-binding as indicated in UniProtKB [77].

The features altered in the SN of female PD patients highlight the importance of acidification, microtubule stability, mitochondrial and lysosomal dysfunction, glutamic metabolism, and neurotoxicity to neurodegeneration and neuronal death in PD. The potential dysfunction of mitochondrial and lysosomal activity in PD remains of particular interest, with both systems playing a vital role in cellular redox homeostasis. Mitochondrial dysfunction can promote a decline in energy production, the increased generation of reactive oxygen species, and the induction of stress-induced apoptosis. Meanwhile, lysosomes participate in the turnover and degradation of organelles and proteins; targets such as the mitochondria and alpha-synuclein aggregates, respectively, may have relevance to PD [78–80].

Notably, the functional profiles inferred in male and female PD patients appear similar (oxidative stress, inflammation, and neurodegeneration); however, we found differences in the protein clusters defined by differential gene expression and the underlying mechanisms. While male PD patients present an environment related to metal homeostasis, lipid metabolism, and immunity, the female PD patients’ environment exhibits mitochondrial and lysosomal dysfunction and alterations to cytoskeletal proteins and glutamic metabolism. Importantly, these data may help further understand PD development and guide personalized, sex-specific therapeutic interventions for PD patients.

Finally, we identified 15 differentially expressed genes in the global meta-analysis. Eleven of these genes appeared significantly increased in male PD patients (AC073283.7, ATP5J2-PTCD1, GPR4, IL15RA, ITGA5, LAMA5, MAP3K6, MEGF6, PNTM, PRDM12, and RASIP1), with the majority associated with oxidative stress, inflammation, and cerebral disorders [81]. The remaining 4 genes displaying a significant increase in female PD patients (OPTN, UBA6, BZW1, and TMX2) have been linked to apoptosis, ubiquitination, and mitochondrial activity. OPTN interacts with adenovirus E3-14.7K protein and mediates apoptosis, inflammation, and vasoconstriction through tumor necrosis factor-alpha or Fas-ligand pathways [82]. Several viruses have been related to the etiology of neural diseases, although the underlying mechanisms remain incompletely understood [83,84]. Ubiquitin dysregulation can affect normal PINK1 and Parkin function, which govern mitochondrial quality control and mitophagy [85]. In female subjects, the alteration of these genes results in mitochondrial dysfunction, stress condition, energy depletion, and necrotic cell death [86,87]. Even though the genes involved differ, we note similar overall results in male and female PD patients: the overrepresentation of mitochondrial dysfunction, oxidative stress, and inflammation, which translates into neuronal death and cognitive/intellectual disorders. This study represents the first description of a sex-based association of these genes in PD, highlighting them as candidates for future studies.

We also note that many genes identified in the SN and global/three-region meta-analysis have been linked to other neural, intellectual, and cognitive disorders in the Open Targets database. In particular, specific genes have been linked to PD, which provides confidence to the meta-analysis results. Further exploration of these genes may open new perspectives for biomarker identification, early diagnostics, and therapeutic approaches in PD and related disorders.

### SN Sex-based Functional Profiling

Based on the differential gene expression analysis in SN, we applied different approaches to analyze the functional scenario. ORA demonstrated significantly more increased genes in male PD patients associated with metal ion detoxification and stress responses to several ions. Detailed studies have encountered metal ions in protein aggregates in PD brains [88,89] and have demonstrated the contribution of metal ions to oxidative stress, toxicity, and degeneration of dopaminergic neurons in PD [90]. In correlation with ORA results, PPI analyses identified metallothioneins, redox homeostasis, oxidative stress, and inflammation as the most prominent protein interaction clusters in male PD patients and angiogenesis. As interferon-stimulated genes [91,92], metallothioneins may combine with Interferon-gamma to upregulate microglia genes, a mechanism observed in multiple system atrophy [93,94]. The MICAL-like protein 2 Ras-related protein Rab is an effector of small Rab GTPs such as Rab13, which modulate alpha-synuclein levels, aggregation, and toxicity [95]. The ARPC1B cluster may be involved in the cytolytic activity of CD8 cytotoxic T lymphocytes [96], which is related to brain inflammation. Several studies support the role of STOM/TMBIM1/CYBA proteins in redox homeostasis and oxidative stress, inflammation, and blood vessel remodeling [77,97]. Finally, increased angiogenesis has been described in SN post-mortem tissues from PD patients [98]

In female PD patients, overrepresented functions correspond to microtubule and cytoskeletal transport, mitochondrial transport along the microtubule, macroautophagy, and several functions associated with antigen processing and presentation. α-synuclein is a primary trigger of the immune response in PD [99] and may activate both the innate and adaptive immune system since the degree of microglial activation directly correlates with α-synuclein load in post-mortem brains [99]. Purine metabolism is also overrepresented in the “LFC > 0” gene set, with levels of urate and other purines known to correlate with PD severity [100]. Regarding purine-ribose dysregulation observed in females, a reported link to microglial activation (neuroinflammation) and neurodegeneration [9] could inspire the design of specific therapeutic strategies in male and female PD patients. As in males, PPI analyses in female PD patients identified clusters closely related to the gene families and ORA results. These clusters provided evidence for the critical role of mitochondria and metabolism in PD, as reported for other neurodegenerative diseases [77]. Glutamate metabolism is vital for neuronal excitability, playing a critical role in memory, synaptic plasticity, and neuronal development [101]; however, glutamate overstimulation is also implicated in toxicity and neurodegeneration [102]. V-ATPase subunits (ATP-dependent proton pumps present in both intracellular compartments and the plasma membrane) have significant involvement in neurodegenerative diseases. Proton pumps contribute to defective lysosomal acidification in lysosomal storage disorders and common neurodegenerative conditions such as Alzheimer’s disease and PD [69]. Supporting our results, the dysregulation of secretogranins in PD has also been reported [67]. Finally, protein folding, recycling, and degradation could be related to the misfolding and aggregation of toxic α-synuclein [103,104].

Overall, the PPI and ORA results correlated well; furthermore, sex-related differences in inflammation, mitochondrial dysfunction, and oxidative stress mechanisms have been documented in several other studies [14,18–20], which provides additional support to our results. Importantly, we identify differences in the protein networks involved in these mechanisms between male and female PD patients, which could be crucial to designing personalized therapeutic strategies.

Transcription factors represent critical modulators of gene expression and pathways controlling responses to stimuli such as oxidative stress, microglial activation, chronic inflammation, neurotoxins, and DNA damage [105]. A wide range of transcription factors have been linked to PD pathogenesis, and as with other cells, the development, maintenance, and survival of neurons depend on the precise control of gene expression [106]. Activated transcription factors in male PD patients included ADNP, JUN, MBD3, PRDM14, and ESR1. In the OpenTargets database [36], alterations in ADNP have been associated with autism spectrum disorders, intellectual disability, dysmorphic features, and hypotonia [107], JUN with Alzheimer’s disease, and PRDM14 with schizophrenia. Meanwhile, MBD3 is a critical neurodevelopmental transcription factor [108,109] associated with neurogenesis and connectivity [108,110]. Alterations in MBD3 have been related to PD [111] and to Neuropathy, Hereditary Sensory, Type Ie. in the GeneCards database [59]. Polymorphisms in ESR1 may contribute to increased PD susceptibility [112]. Male PD patients also displayed the altered activity of SP1, which regulates the expression of LRRK2 [113], a contributing factor to PD pathogenesis; furthermore, a study has suggested that SP1 inhibition may provide beneficial effects in PD models [114]. SMAD3 also presents higher activity in male PD patients and plays a vital role in PD, with protein deficiency known to reduce neurogenesis significantly. SMAD3 dysfunction leads to the formation of α-synuclein aggregates and a reduction in the number of dopaminergic axons and dendrites [115]. Other transcription factors altered in male PD patients (i.e., BATF, CEBPA, ETS1, KLF6, LYL1, NFKB1, RELA, SOX13, SP3, SPI1, STAT1, STAT2, and STAT3) may participate in neuroinflammatory processes and regulate various functions of the immune response.

Of the transcription factors altered in female PD patients, ATF3, BCL6, and PCGF2 have been previously associated with neurodegenerative diseases or cognitive disabilities. Alterations in ATF3 in response to neurological damage and reactive oxygen species production have been demonstrated in a PD model [116,117]. The BCL6 transcriptional repressor targets the ITM2B gene [118], which has important links to neurodegenerative diseases such as AD. The ITM2B protein binds amyloid precursor protein and inhibits processing, thereby reducing the secretion and accumulation of beta-amyloid peptides. PCGF2 is associated with phenotypes such as intellectual disability, global developmental delay, and mental retardation in the OpenTargets database [36]. Additionally, the ZBTB7A, ZC3H8, or NME2 transcription factors have been linked to neuroinflammation processes. These transcription factors play roles in multiple processes, including the immune response, modulating hemopoiesis, and regulating the differentiation and activation of T and B lymphocytes (Additional file 3: Table S3). The transcription factors identified in this study may represent biomarkers for the detection of neuron degeneration, although this will require more in-depth validation studies.

### Strengths and Limitations

We performed an *in-silico* approach to study sex-based differences in PD. *In-silico* strategies using computational models represent a powerful tool to evaluate and integrate data, and the results obtained possess greater consensus and statistical power. In this approach, the sample size increases with an augmentation of the number of studies integrated into the meta-analysis; therefore, more subtle effects can be detected. Additionally, we based our *in-silico* analysis of PD with a sex perspective on FAIR data (Findable, Accessible, Interoperable, Reusable) [119], which we believe to be critically important. Indeed, we strongly believe that research data should be legally sharable and reusable in new research. *In-silico* integrative approaches to analyzing sex-based differences in PD have been carried out before; for example, Mariani et al. [120] systematically meta-analyzed SN microarray data using the Transcriptome Mapper software (TRAM version 1.2). Our study also includes 2 other primarily affected brain regions (FC and ST) and has been carried out using the limma R package for differential expression analyses [30] and the metafor R package for gene expression meta-analysis [33]. This allowed us to analyze data following our sex differences comparison, which includes 4 experimental groups, and identifies sex differences in PD, considering the inherent variability among males and females in healthy conditions.

Sex-based differences influence progression, symptoms, treatment response, and mortality in many diseases, including PD; however, the segregation of data by sex does not always occur in research studies, not even in those carried out to explore diagnostic or prognostic factors. For example, we excluded 44 (53%) studies from our systematic review due to the absence of sex information. We highlight the need to include sex information in research studies and databases, as they are relevant to health.

## CONCLUSIONS

In conclusion, our *in-silico* approach has highlighted sex-based differential mechanisms in typical PD hallmarks (inflammation, mitochondrial dysfunction, and oxidative stress). Additionally, we have identified specific genes and transcription factors for male and female PD patients that represent potential candidates as biomarkers to diagnosis. Further studies that consider the sex perspective are urgently required to better understand PD and develop tailored interventions that consider the distinct requirements of male and female PD patients. Finally, we underscore the essential nature of sharing data and using open platforms for scientific advancement.

## Data Availability

DATA AVAILABILITY. The data used for the analyses described in this work is publicly available at GEO (https://www.ncbi.nlm.nih.gov/geo/) and ArrayExpress (https://www.ebi.ac.uk/arrayexpress/). The accession numbers of the GEO datasets downloaded are: GSE8397, GSE20295, GSE7621, GSE20146 and GSE28894. The accession numbers of the ArrayExpress datasets downloaded are: E-MTAB-1194 and E-MEXP-1416.

https://github.com/adlpecer/PD_GeneMetaanalysis

http://bioinfo.cipf.es/metafun-pd

## ADDITIONAL FILES

**Additional file 1: Table S1**. Results for all significant genes from all meta-analyses. It includes the adjusted p-value, the LFC and its 95% confidence interval (CI), and the standard error (SE) of the LFC. (XLSX 26 kB)

**Additional file 2: Table S2**. List of R packages and versions used in this work. (XLSX 9 kB)

**Additional file 3: Table S3**. VIPER results for each TF evaluated. It includes the adjusted p-value, the NES, the size of the regulons and their functional annotation (From Gene Ontology Biological Processes). (XLSX 24 kB)

### ABBREVIATIONS

FC: Frontal cortex
GEO: Gene Expression Omnibus
LFC: Log_2_ fold change
NES: Normalized enrichment score
ORA: Over-representation analysis
PD: Parkinson’s disease
PPI: Protein-protein interaction
PRISMA: Preferred reporting items for systematic reviews and meta-analyses
SN: Substantia nigra
ST: Striatum

## DATA AVAILABILITY

The data used for the analyses described in this work are publicly available at GEO [23] and ArrayExpress [24]. The accession numbers of the GEO datasets downloaded are GSE7621, GSE8397, GSE20146, GSE20295, and GSE28894. The accession numbers of the ArrayExpress datasets downloaded are E-MTAB-1194 and E-MEXP-1416.

## CODE AVAILABILITY

The code developed for the analyses described in this work has been made publicly available at GitHub (https://github.com/adlpecer/PD_GeneMetaanalysis). All software and versions used are detailed in Additional file 2: Table S2.

## ACKNOWLEDGEMENTS

The authors thank the Principe Felipe Research Center (CIPF) for providing access to the cluster, which is co-funded by European Regional Development Funds (FEDER) in Valencian Community 2014-2020. This work was supported by GV/2020/186. The authors also thank Stuart P. Atkinson for reviewing the manuscript.

## AUTHOR CONTRIBUTIONS

Conceptualization, F.G.G.; methodology, A.L.C., Z.A., R.G.R., M.R.H., and F.G.G.; software, A.L.C., and M.R.H.; validation, A.L.C., M.R.H., R.G.R., and F.G.G.; formal analysis, JF.C.S., R.G.R., and A.L.C.; investigation, A.L.C., Z.A., and F.G.G.; data curation, A.L.C., and M.R.H.; writing—original draft preparation, A.L.C., Z.A., M.R.H., R.G.R., JF.C.S., I.S.S., A.N.A., M.I.V., F.G., and F.G.G.; writing—review and editing, A.L.C., Z.A., M.R.H., R.G.R., JF.C.S., I.S.S., A.N.A., M.I.V., F.G., and F.G.G.; visualization, A.L.C., R.G.R., M.R.H., and F.G.G.; supervision, Z.A., M.R.H., and F.G.G.; funding acquisition, F.G.G.; project administration, F.G.G. All authors have read and agreed to the published version of the manuscript.

## COMPETING INTERESTS

The authors declare no competing interests.

**Correspondence** and requests for materials should be addressed to F.G.G.

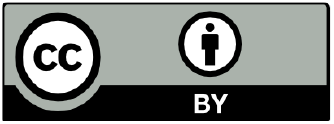

**Open Access** This article is licensed under a Creative Commons Attribution 4.0 International License, which permits use, sharing, adaptation, distribution and reproduction in any medium or format, as long as you give appropriate credit to the original author(s) and the source, provide a link to the Creative

Commons license, and indicate if changes were made. The images or other third-party material in this article are included in the article’s Creative Commons license, unless indicated otherwise in a credit line to the material. If material is not included in the article’s Creative Commons license and your intended use is not permitted by statutory regulation or exceeds the permitted use, you will need to obtain permission directly from the copyright holder. To view a copy of this license, visit http://creativecommons.org/licenses/by/4.0/

## Notes

### Competing Interest Statement

The authors have declared no competing interest.

### Author Declarations

Gene Expression Omnibus (GEO) and ArrayExpress databases

### Summary of Updates

We have updated some sections

